# Cross-reactive antibody and functional T-cell responses to Bundibugyo virus induced by Zaire ebolavirus vaccination and infection

**DOI:** 10.64898/2026.11.03.26367168

**Authors:** Aurelie Wiedemann, Ahidjo Ayouba, Edouard Lhomme, Emile Foucat, Solenne Kisrani, Guillaume Thaurignac, Safaa Ben-Farhat, Postebogui Study group, Prevac study team, Martine Peeters, Yves Lévy

## Abstract

**Background:** There is no licensed specific vaccine against the current Ebola Bundibugyo virus (BDBV) outbeak affecting the Democratic Republic of the Congo. However, clinical evaluations of a licensed Zaire ebolavirus (EBOV) vaccine have just launched. It remains unclear whether EBOV infection or vaccination generate cross-reactive immunity against BDBV.

**Methods:** We quantified CD4⁺ and CD8⁺ T-cell magnitude, polyfunctionality, and cytotoxicity after 8-day expansion with EBOV, BDBV, Sudan (SUDV), or Marburg (MARV) GP peptide pools, and measured GP-binding antibodies by multiplex assay, in recipients of Ad26.ZEBOV/MVA-BN-Filo (n=10), single-dose rVSVΔG-ZEBOV-GP (n=10), or two-dose rVSVΔG-ZEBOV-GP (n=8) (NCT02876328), and in EVD survivors sampled approximately 3 years after infection (n=10).

**Findings:** Fourteen days after completion of each licensed vaccine regimen, median BDBV-reactive CD4⁺ responses were 9.8% [IQR:8.9-14.0] after single-dose rVSVΔG-ZEBOV-GP (D14) and 8.0% [5.5-9.9] after Ad26.ZEBOV/MVA-BN-Filo (D70). The highest BDBV-reactive CD8⁺ response was observed in survivors (24.7% [20.3-33.1]), together with a prominent cytotoxic CD8⁺ response (14.4% [10.4-19.7]); corresponding vaccine-group medians ranged from 0.9% to 17.7% and from 0.6% to 6.7%, respectively. SUDV responses were also detected, whereas MARV recognition was limited. BDBV-binding antibodies were detected following both vaccine regimens, with the highest levels observed in survivors (491 [330-1261] net MFI). Among vaccinees, EBOV- and BDBV-reactive cellular and humoral responses were positively correlated after adjustment for multiple testing.

**Interpretation:** EBOV infection and vaccination elicited BDBV-cross-reactive T-cell responses and binding antibodies. These findings support further evaluation of existing EBOV vaccines in the context of BDBV outbreaks.

## Introduction

By Oct 27, 2026, the Bundibugyo virus (BDBV) outbreak in the Democratic Republic of the Congo had surpassed 10 000 laboratory-confirmed cases and spread regionally, highlighting the urgent need for countermeasures against non-Zaire ebolaviruses ^1^ ^2^. No BDBV-specific vaccine is currently licensed. rVSVΔG-ZEBOV-GP remains licensed for the prevention of EBOV disease, whereas the EU marketing authorisations for Ad26.ZEBOV and MVA-BN-Filo were withdrawn in May, 2026, at the manufacturer’s request for commercial reasons. However, in the absence of established clinical efficacy against BDBV, additional immunological data regarding the licensed EBOV vaccines are critically required to evaluate potential heterologous protection and support their evidence-based deployment against the ongoing BDBV outbreak.

In non-human primates, rVSVΔG-ZEBOV-GP provided partial protection against heterologous BDBV challenge ^3^, but protection was not consistently demonstrated across experimental models ^4^. Although no correlates of protection against Ebola viruses were identified, survival following experimental BDBV infection was associated with early antibody responses and activation of cytotoxic T-cell and other adaptive immune pathways, suggesting that both humoral and cellular immunity may contribute to protection ^5^.

We showed that both natural EBOV infection and EBOV vaccination induce durable, polyfunctional memory T-cell responses ^6,7^. Nevertheless, the extent of cross-reactive immunity induced by EBOV vaccination or natural infection remains uncertain. Experimental studies suggest that the magnitude of antibody and T-cell cross-reactivity follows filovirus phylogenetic relatedness ^8^. While a study reported predominantly species-specific antibody responses following Ad26.ZEBOV/MVA-BN-Filo vaccination or BDBV infection in humans ^9^, we recently showed cross binding BDBV antibodies following both rVSVΔG-ZEBOV-GP and Ad26.ZEBOV/MVA-BN-Filo vaccination ^10^.BDBV-reactive T cells have recently been detected following rVSVΔG-ZEBOV-GP vaccination ^11^. However, the breadth, lineage distribution and functional quality of these responses across vaccine strategies and following natural EBOV infection remain poorly defined, in particular their polyfunctional and cytotoxic properties. Furthermore, whether different EBOV vaccine strategies and natural infection generate comparable patterns of heterologous recognition remains unknown.

Here, we analysed paired serum and peripheral blood mononuclear cell samples from healthy participants enrolled in the PREVAC vaccine trial ^7^ and Ebola virus disease (EVD) survivors from the Postebogui cohort ^6^ to characterize both humoral and cellular responses to EBOV, BDBV, SUDV and MARV. We assessed the magnitude, relative breadth, polyfunctionality and cytotoxic potential of heterologous CD4⁺ and CD8⁺ T-cell responses and characterized filovirus GP-binding antibody responses in the same study populations. This integrated analysis provides evidence on the breadth and functional quality of immunity induced by EBOV vaccination and natural infection in humans and may inform the evaluation of existing EBOV vaccines during the ongoing BDBV outbreak.

## Methods

### Study participants and samples

Participants were selected from two previously described studies: the multicentre, randomised, placebo-controlled phase II PREVAC vaccine trial (NCT02876328) and the Postebogui cohort of laboratory-confirmed EVD survivors, both conducted in Guinea ^12–16^.

From PREVAC, adult participants enrolled in Guinea were randomised to Ad26.ZEBOV followed by MVA-BN-Filo 56 days later (Ad26-MVA), rVSVΔG-ZEBOV-GP followed by placebo (rVSV), rVSVΔG-ZEBOV-GP followed by a homologous booster (rVSV-booster), or placebo. Samples were randomly selected based on residual PBMC and serum availability, independently of previous immunological results. PBMC samples were available at baseline (D0; n=5), 14 days after priming (D14; n=20: rVSV, n=10; Ad26.ZEBOV, n=10), and 14 days after the second vaccination (D70; n=18: rVSV–booster, n=8; Ad26-MVA, n=10). Serum samples were available at D0 (n=5), D7 (n=28), D63 (n=18), and M3 (n=28).

From Postebogui, PBMC and serum samples were obtained from 10 EVD survivors approximately 3 years after discharge from an Ebola treatment centre. All participants provided written informed consent for sample collection, storage, and use.

Study protocols were approved by the relevant ethics committees in Guinea and France, including INSERM/CEEI and IRD/CCDE for Postebogui and INSERM IRB00003888 for PREVAC. PREVAC was registered with ClinicalTrials.gov (NCT02876328), EudraCT (2017-001798-18), and PACTR (PACTR201712002760250). Sample processing and storage followed previously described procedures ^6,7^.

### Characterization of EBOV-specific immune responses

Cellular responses to filovirus GP peptide pools were assessed using the EpiMax assay, as previously described ^6,7^. Briefly, PBMCs were expanded in vitro with pools of overlapping 15-mer peptides with an 11-amino-acid overlap spanning the full-length GP of Ebola virus Mayinga (167 peptides), Bundibugyo virus Uganda 2007 (167 peptides), Sudan virus Uganda 2000 (167 peptides), or Marburg virus Musoke 1980 (168 peptides) (JPT Peptide Technologies, Berlin, Germany). T-cell responses and functional profiles were subsequently assessed by intracellular cytokine staining and flow cytometry. The panel included a viability marker and antibodies against CD3, CD4 and CD8 for T-cell lineage identification, together with CD107a, IFN-γ, TNF, IL-2 and granzyme B for functional characterization. Polyfunctionality was evaluated using Boolean combinations of IFN-γ, TNF and IL-2 expression, while cytotoxic T-cell responses were defined based on CD107a and granzyme B co-expression. Functional profiles were visualized using SPICE version 5.22.

Cross-reactivity ratios were calculated separately for CD4⁺ and CD8⁺ T cells as the percentage of background-subtracted heterologous (BDBV, SUDV or MARV) responses relative to the corresponding EBOV response, at the individual participant level. Ratios were calculated only for EBOV responders (antigen-stimulated responses ≥3-fold above background). Negative background-subtracted heterologous responses were set to zero, and ratios exceeding 100% were retained. Data are presented as individual values with medians and interquartile ranges. Absolute responses for all participants, including EBOV non-responders, are shown in Figure 1.

**Figure 1.**
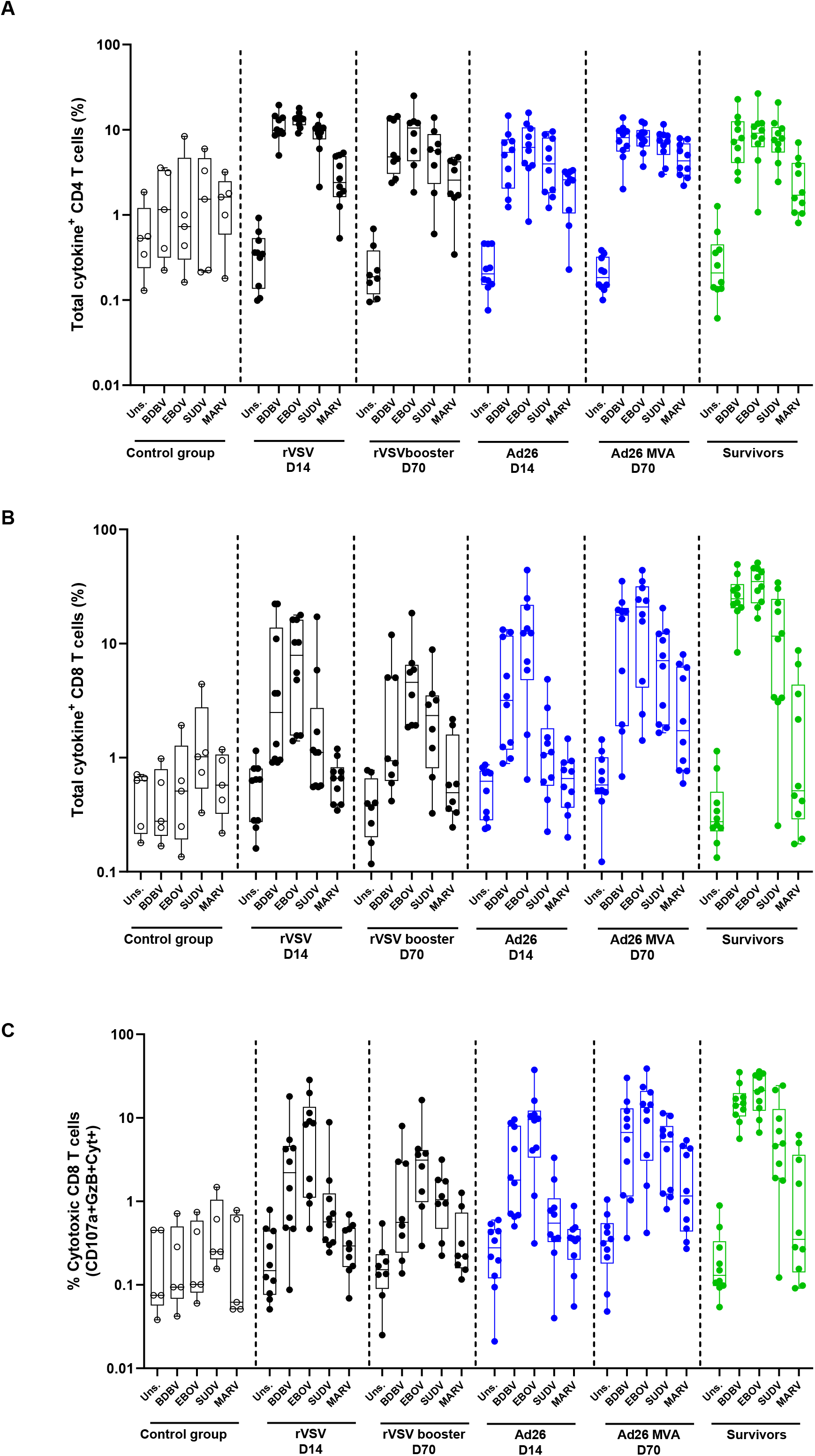
Magnitude and cytotoxic profile of filovirus GP-specific T-cell responses following EBOV vaccination and natural infection. PBMCs from healthy unvaccinated participants (control group (collected before vaccination); n=5), recipients of a single dose of rVSVΔG-ZEBOV-GP assessed at D14 (n=10), recipients of two doses of rVSVΔG-ZEBOV-GP assessed at D70 (n=8), recipients of Ad26.ZEBOV assessed at D14 (n=10), recipients of Ad26.ZEBOV/MVA-BN-Filo assessed at D70 (n=10), and EVD survivors sampled approximately three years after discharge from an Ebola treatment centre (n=10) were stimulated with overlapping GP peptide pools from BDBV, EBOV, SUDV or MARV following antigen-specific 8-day in vitro expansion. Unstimulated cultures (Uns.) were included as negative controls. Frequencies of total cytokine-producing (A) CD4⁺ and (B) CD8⁺ T cells, defined by the expression of IFN-γ, IL-2 and/or TNF, are shown as percentages of total CD4⁺ or CD8⁺ T cells, respectively. (C) Filovirus-specific CD8⁺ T cells with a cytotoxic phenotype were defined by the co-expression of CD107a and granzyme B together with at least one cytokine (IFN-γ, IL-2 and/or TNF) and are shown as percentages of total CD8⁺ T cells. Each symbol represents an individual participant. Boxes indicate the median and interquartile range, and whiskers represent min-max values.

### Multiplex assessment of cross-reactive antibody responses

Cross-reactive antibody responses were assessed using a multiplex bead-based Luminex assay. Recombinant glycoproteins from EBOV Mayinga, EBOV Kikwit, BDBV Uganda 2007, SUDV Gulu, and MARV Musoke 1980 were coupled to distinct bead regions. Serum samples were tested in duplicate at a 1:200 dilution and incubated with antigen-coated beads for 16 h at 4°C under agitation and protected from light. Bound antibodies were detected using biotinylated mouse anti-human immunoglobulin G antibodies followed by streptavidin–phycoerythrin. Median fluorescence intensity was measured using a Luminex INTELLIFLEX platform. Samples with a coefficient of variation greater than 20% between duplicates were retested, and the median value of duplicate measurements was used for statistical analyses ^17^.

### Statistical analysis

Magnitude and cytotoxic profile of filovirus GP-specific T-cell responses following EBOV vaccination and natural infection, and Filovirus GP-specific antibody-binding responses following EBOV vaccination and natural infection were described in terms of median and interquartile range. Associations between EBOV- and BDBV-specific responses were assessed using Spearman’s rank correlation coefficient. Analyses were performed separately in vaccinees and EBOV survivors. To account for multiple testing, p-values were adjusted using the Benjamini–Hochberg false discovery rate procedure within the humoral and cellular response analyses, considered as distinct families of tests. Analyses were performed using R (the R Foundation, Vienna, Austria; version 4.2) software.

## Results

### Cross-reactive memory T-cell responses in EBOV vaccine recipients and EVD survivors

To characterize the breadth of memory T-cell responses induced by EBOV vaccination or infection, we assessed CD4⁺ and CD8⁺ T-cell responses to EBOV, BDBV, SUDV and MARV GP peptide pools following antigen-specific in vitro expansion. Analyses included samples collected before vaccination (D0; n=5), 14 days after a single dose of rVSVΔG-ZEBOV-GP (D14; n=10) or Ad26.ZEBOV (D14; n=10), and 14 days after boosting at D56 with rVSVΔG-ZEBOV-GP (D70; n=8) or MVA-BN-Filo (D70; n=10). Samples from EVD survivors of the 2014 EBOV outbreak collected approximately three years after discharge from an Ebola treatment center (n=10) were included (**Figure 1**).

In participants sampled before vaccination, cytokine-producing CD4⁺ T cells, defined by the expression of IFN-γ, IL-2 and/or TNF, were detected at low frequencies following stimulation with each of the four filovirus GP peptide pools. Median frequencies [IQR] were 1.1% [0.3-3.3] for BDBV, 0.7% [0.3-4.7] for EBOV, 1.5% [0.2-4.6] for SUDV and 1.6% [0.6-2.5] for MARV, compared with 0.5% [0.2-1.2] in unstimulated cultures (**Figure 1A**).

Following a single dose of rVSVΔG-ZEBOV-GP, broad CD4⁺ T-cell responses were detected against both homologous and heterologous filovirus antigens. The highest median frequency was observed following EBOV stimulation (13.4% [11.3–14.2]), while responses to BDBV and SUDV reached 9.8% [8.9-14.0] and 9.7% [7.7-10.7], respectively. The median response to MARV was lower, at 2.4% [1.6-4.9], compared with 0.3% [0.1-0.5] in unstimulated cultures. Response magnitudes following the second rVSVΔG-ZEBOV-GP dose remained within the same range as those observed after the first dose.

Broad CD4⁺ T-cell responses were also observed following the Ad26.ZEBOV/MVA-BN-Filo regimen. Responses to all four peptide pools were detectable after Ad26.ZEBOV priming and remained detectable following MVA-BN-Filo administration. At D70, median frequencies were 8.0% [5.5-9.9] for BDBV, 8.2% [6.4-9.9] for EBOV, 7.4% [5-8.7] for SUDV and 4.3% [2.9-6.9] for MARV, compared with 0.2% [0.1-0.3] in unstimulated cultures. Of note, because MVA-BN-Filo encodes the glycoproteins of EBOV, SUDV and MARV, responses to SUDV and MARV at D70 cannot be attributed to cross-reactivity alone.

Broad CD4⁺ T-cell recognition was similarly detected in EVD survivors approximately three years after discharge from an Ebola treatment centre. Median responses to BDBV, EBOV and SUDV were within the same range, reaching 7.6% [4-12.5], 9.1% [6.2-11.8] and 7.7% [5.4-10.7], respectively. The median response to MARV was 1.7% [1.0-4.0], compared with 0.2% [0.1-0.4] in unstimulated cultures (**Figure 1A**).

Overall, the descriptive response profiles observed after EBOV vaccination and infection were characterized by broad CD4⁺ T-cell recognition of EBOV, BDBV and SUDV antigens, with lower responses to MARV.

CD8⁺ T-cell responses showed an overall pattern similar to that observed for CD4⁺ T cells, with broad recognition of EBOV, BDBV and SUDV following vaccination and natural infection, and consistently lower responses to MARV. Responses were detected after a single dose of either rVSVΔG-ZEBOV-GP or Ad26.ZEBOV and remained detectable following the second vaccination. Notably, the strongest CD8⁺ T-cell responses to EBOV, BDBV and SUDV peptide pools were observed in EVD survivors where median CD8⁺ T-cell frequencies reached 34.9% [22.6-45.4] for EBOV, 24.7% [20.3-33.1] for BDBV and 11.6% [3.3-24.6] for SUDV, compared with 0.5% [0.3-4.3] for MARV (**Figure 1B**).

We next assessed the cytotoxic potential of filovirus-specific CD8⁺ T cells based on the co-expression of CD107a and granzyme B. Cytotoxic CD8⁺ T-cell responses were detected against EBOV, BDBV and SUDV across vaccine groups and in EVD survivors, whereas responses to MARV remained limited. The most prominent responses to EBOV and BDBV were observed in survivors, with median frequencies of 21.2% [12.1-32.6] and 14.4% [10.4-19.7], respectively, exceeding the median responses observed across vaccine groups (range from 3.1 to 13.3% for EBOV, and from 0.6 to 6.7% for BDBV). Responses to SUDV were also readily detectable in survivors (4.8% [1.9-12.7]), whereas MARV-specific responses remained low (0.4% [0.1-3.6]) (**Figure 1C**). These findings reveal a particularly strong cytotoxic CD8⁺ T-cell memory response to both homologous EBOV and heterologous BDBV following natural EBOV infection.

We next examined whether recognition of heterologous filovirus antigens was associated with distinct functional profiles. Boolean analysis of IFN-γ, IL-2 and TNF expression revealed broadly similar CD4⁺ T-cell response profiles across EBOV, BDBV, SUDV and MARV peptide stimulation and among vaccine groups and EVD survivors (**Supplementary Figure 1A**). Responses were predominantly composed of IFN-γ-producing cells, with smaller proportions of cells co-expressing two or three cytokines. CD8⁺ T-cell responses to EBOV, BDBV and SUDV showed a similarly conserved functional composition across groups, dominated by IFN-γ-producing cells and accompanied by smaller polyfunctional subsets (**Supplementary Figure 1B**). MARV-specific CD8⁺ T-cell responses were not included in the polyfunctionality analysis because too few participants in each group met the predefined response criterion of a stimulated response at least threefold higher than the corresponding unstimulated background. Overall, cross-reactive recognition of BDBV and SUDV was not associated with a major shift in the functional composition of the T-cell response relative to EBOV.

We then assessed the relative breadth of T-cell recognition by expressing heterologous responses as percentages of the corresponding EBOV response. For CD4⁺ T cells, BDBV and SUDV responses represented substantial proportions of EBOV-specific responses across groups, with median ratios ranging from 71.2% to 93.2% and 51.5% to 90.2%, respectively. The highest ratios were observed in EVD survivors (88.6% for BDBV and 90.2% for SUDV) and Ad26-MVA vaccinee at D70 (93.2% for BDBV and 85.5% for SUDV). In contrast, MARV recognition was generally limited (13.6-20.9% after rVSVΔG-ZEBOV-GP vaccination and in survivors, but higher following Ad26.ZEBOV priming and MVA-BN-Filo administration (**Supplementary Figure 2A**).

CD8⁺ T-cell ratios were more heterogeneous. BDBV responses represented 23.7 to 64.8% of EBOV responses across vaccine groups, reaching 84.0% [55.0-101.9] in EVD survivors. SUDV recognition varied considerably, from 0.4% after Ad26.ZEBOV priming to 52.6% following MVA-BN-Filo administration and 43.3% in survivors. MARV recognition remained minimal in most groups, with higher relative responses after MVA-BN-Filo administration (11.2% [0.3-22.9]). The higher SUDV and MARV ratios following MVA-BN-Filo administration were consistent with its multivalent antigenic composition (**Supplementary Figure 2B**).

Together, these findings demonstrate broad BDBV-reactive CD4⁺ and CD8⁺ T-cell responses following EBOV vaccination and natural infection. Cross-reactive recognition was more consistently preserved within the CD4⁺ compartment, whereas CD8⁺ T-cell cross-reactivity was more heterogeneous but remained particularly prominent against BDBV, especially in EBOV survivors.

### Cross-reactive antibody responses in vaccine recipients and EVD survivors

To assess whether heterologous T-cell recognition was accompanied by humoral responses, we measured serum IgG binding to BDBV, EBOV Mayinga and Kikwit, SUDV, and MARV glycoproteins using a multiplex bead-based assay at D0, D7, D63, M3, and in EVD survivors.

At D7, antibody-binding levels following either rVSVΔG-ZEBOV-GP or Ad26.ZEBOV priming remained comparable to those in unvaccinated controls. Following the second vaccination, strong binding to both EBOV variants was observed, with median [IQR] net MFI values of 5059 [3548-8971] and 3436 [1873-8019] for Mayinga, and 2920 [2515-4179] and 1390 [737-3193] for Kikwit, in the rVSV and Ad26/MVA groups, respectively (**Figure 2A**). BDBV binding was more modest, with median levels of 238 [201-284] and 202 [102-775] net MFI at D63, and 157 [86-280] and 243 [160-447] at M3, respectively (**Figure 2A,B**). SUDV-binding levels largely overlapped with those of unvaccinated controls, while MARV binding remained low, including after MVA-BN-Filo administration. Notably, responses to SUDV and MARV following MVA-BN-Filo cannot be attributed to cross-reactivity alone, as this vaccine encodes both glycoproteins (**Figure 2B**).

**Figure 2.**
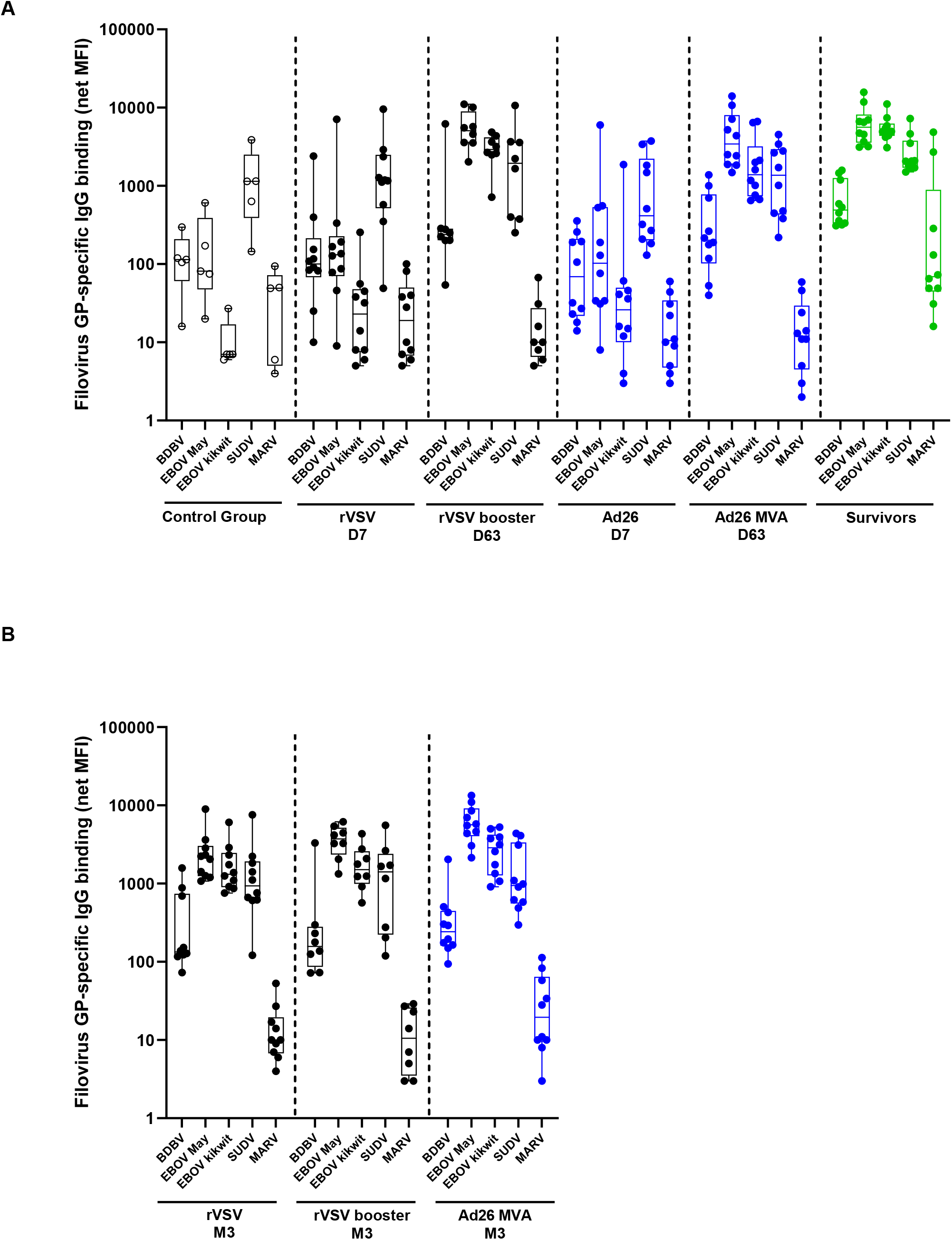
Filovirus GP-specific antibody-binding responses following EBOV vaccination and natural infection. Serum IgG binding to recombinant glycoproteins from BDBV, two EBOV variants (Mayinga and Kikwit), SUDV and MARV was measured using a multiplex bead-based assay. (A) Early antibody-binding responses were assessed in healthy unvaccinated participants (control group, collected before vaccination n=5), recipients of rVSVΔG-ZEBOV-GP seven days after the first vaccination (D7, n=10), recipients of two doses of rVSVΔG-ZEBOV-GP seven days after the second vaccination (D63, n=8), recipients of Ad26.ZEBOV at D7 (n=10), recipients of Ad26.ZEBOV/MVA-BN-Filo at D63 (n=10), and EBOV survivors (n=10) sampled approximately three years after discharge from an Ebola treatment centre. (B) Antibody-binding responses at month 3 (M3) were assessed in recipients of a single dose of rVSVΔG-ZEBOV-GP (n=10), two doses of rVSVΔG-ZEBOV-GP (n=8), or the Ad26.ZEBOV/MVA-BN-Filo regimen (n=10). Antibody-binding levels are expressed as background-subtracted median fluorescence intensity (net MFI). Each symbol represents an individual participant. Boxes indicate the median and interquartile range, and whiskers represent min-max values.

EVD survivors exhibited high binding antibody levels to EBOV Mayinga and Kikwit (5620 [3559-8182] and 4950 [4349-6248] net MFI, respectively), together with detectable BDBV (491 [330-1261]) and SUDV (2063 [1702-3791]) binding, whereas MARV binding remained lower (69 [44-890] net MFI; **Figure 2A**).

Overall, high EBOV-binding antibody levels after the second vaccination and among EVD survivors were accompanied by detectable BDBV binding.

### Association between EBOV- and BDBV-specific immune responses

We next examined whether the magnitude of EBOV-specific responses was associated with BDBV recognition within the cellular and humoral compartments. In exploratory analyses pooling vaccine groups, EBOV- and BDBV-reactive total cytokine-producing CD4⁺ T-cell responses were strongly correlated at both D14 (Spearman ρ=0.93, adjusted p=7.49×10⁻⁸) and D70 (ρ=0.76, adjusted p=0.0024). Positive correlations were also observed for CD8⁺ T-cell responses at D14 (ρ=0.57, adjusted p=0.033) and D70 (ρ=0.88, adjusted p=3.00×10⁻⁵); (**Figure 3A**). At Month 3, BDBV GP-binding antibody levels were positively correlated with binding to both EBOV Mayinga GP (ρ=0.71, adjusted p=3.73×10⁻⁵) and EBOV Kikwit GP (ρ=0.46, adjusted p=0.014) (**Figure 3B**).

**Figure 3.**
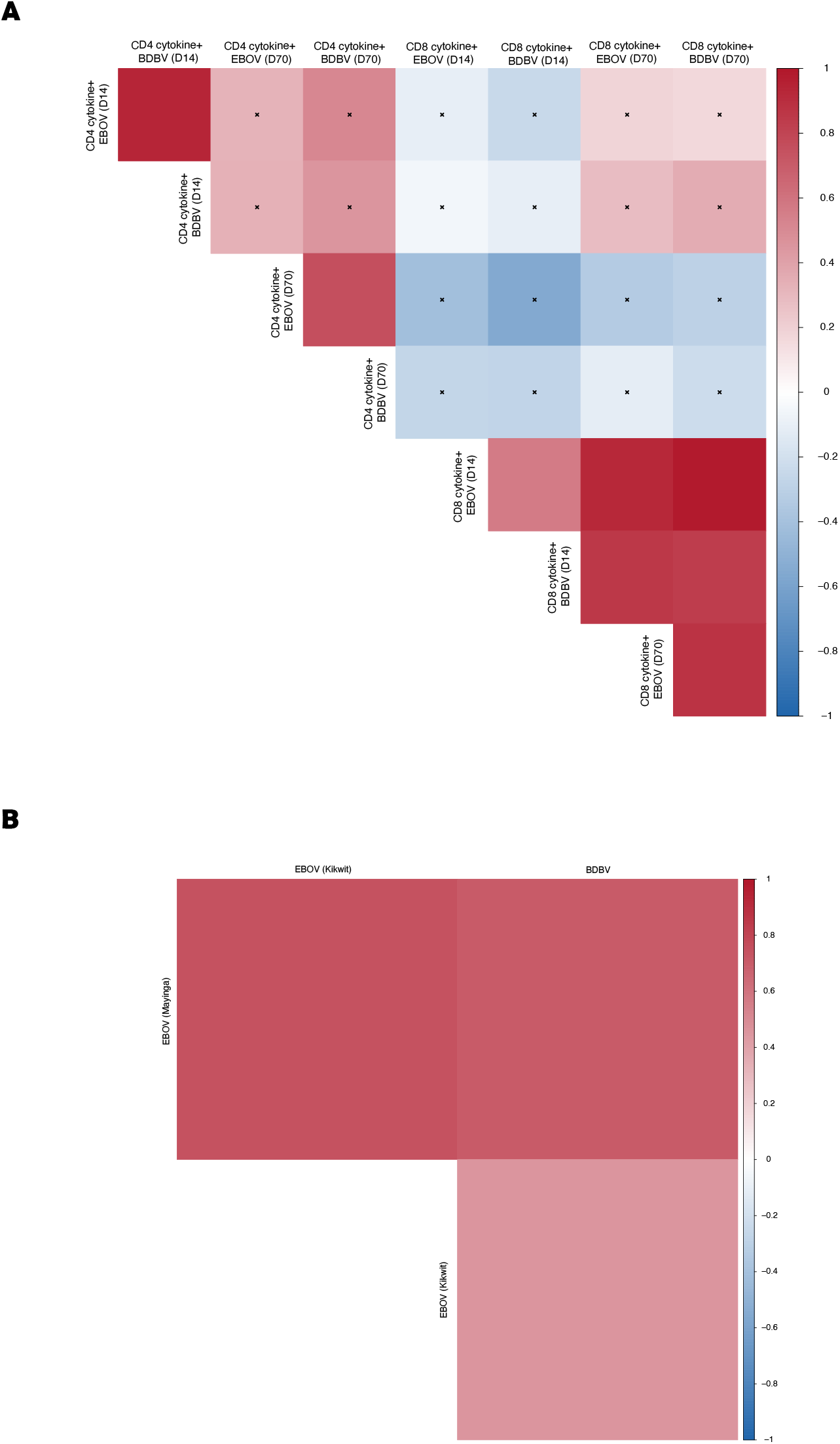
Correlations between EBOV- and BDBV-reactive immune responses in vaccine recipients. (A) Spearman correlation matrix of total cytokine-producing CD4⁺ and CD8⁺ T-cell responses to EBOV and BDBV GP peptide pools at days 14 and 70. Total cytokine-producing cells were defined as cells expressing IFN-γ, IL-2 and/or TNF. (B) Spearman correlation matrix of antibody-binding responses to EBOV Mayinga, EBOV Kikwit and BDBV glycoproteins at Month 3. Vaccine groups were pooled separately at each timepoint (n=20 at D14, n=18 at D70, and n=28 at M3). Colours indicate Spearman correlation coefficients, ranging from −1 (blue) to +1 (red). Crosses indicate correlations that were not statistically significant after adjustment for multiple testing (adjusted P ≥ 0.05).

In EVD survivors, EBOV- and BDBV-reactive CD4⁺ T-cell responses were positively correlated (Spearman ρ=0.81, adjusted p=0.029), whereas the corresponding CD8⁺ T-cell association did not reach statistical significance (ρ=0.65, adjusted p=0.128). Similarly, BDBV GP-binding antibody levels were positively correlated with binding to EBOV Mayinga GP (ρ=0.67, adjusted p=0.0496), whereas the association with EBOV Kikwit GP was not statistically significant (ρ=0.49, adjusted p=0.15) (**Supplementary Figure 3**).

Overall, the analyses indicate that the magnitude of cellular and humoral responses responses against EBOV and heterologous BDBV GPs following vaccination are significantly correlated.

## Discussion

In this study, we provide an integrated characterization of cellular and humoral recognition of BDBV, SUDV and MARV following EBOV vaccination or natural infection. Both vaccine strategies and natural EBOV infection were associated with heterologous recognition of BDBV, involving CD4⁺ and CD8⁺ T-cell responses as well as antibody binding. SUDV was also broadly recognized, although interpretation of vaccine-induced cross-reactivity was complicated by baseline antibody binding and by the inclusion of SUDV GP in MVA-BN-Filo. In contrast, recognition of the more distantly related MARV was consistently limited. Both vaccine strategies and previous EBOV infection generated memory T cells capable of recognizing BDBV and SUDV GP peptide pools, whereas recognition of the more distantly related MARV was consistently limited. Heterologous responses involved both CD4⁺ and CD8⁺ T cells and included polyfunctional and cytotoxic populations. EVD survivors displayed a similar recognition hierarchy but with higher magnitude of the responses than vaccinees, particularly within the cytotoxic CD8⁺ T-cell compartment. These cellular responses were accompanied by antibody binding to BDBV and SUDV, supporting broad, although quantitatively heterogeneous, immune recognition across related orthoebolaviruses.

Our findings extend recent evidence that rVSVΔG-ZEBOV-GP vaccination induces BDBV-reactive T-cell responses in humans. Mayer and colleagues detected BDBV GP-reactive IFN-γ responses in 13 of 20 vaccine recipients at day 28, compared with SUDV-reactive responses in seven participants, and identified recognition of several conserved GP regions ^11^. Our study extends these observations by separately characterizing CD4⁺ and CD8⁺ T-cell responses, assessing their polyfunctional and cytotoxic properties, and comparing rVSVΔG-ZEBOV-GP vaccination with the Ad26.ZEBOV/MVA-BN-Filo regimen and approximately 3 years in survivors of acute EBOV infection. The availability of cellular and humoral measurements further allowed us to examine cross-reactive immunity across complementary immune compartments.

The detection of cross-reactive cytotoxic CD8⁺ T cells may be biologically relevant, although their contribution to protection is likely to depend on the vaccine platform. In non-human primates vaccinated with a recombinant adenovirus expressing EBOV GP, depletion of CD8⁺ cells largely abrogated protection against lethal EBOV challenge, demonstrating a major contribution of cellular immunity ^18^. By contrast, following rVSVΔG-ZEBOV-GP vaccination, CD8⁺ T-cell depletion did not compromise survival, whereas depletion of CD4⁺ T cells during vaccination impaired the induction of GP-specific antibodies and abrogated protection, supporting a predominant role for antibody-mediated immunity in this model ^19^. Indeed, although vaccine-induced EBOV GP-binding antibodies are considered a putative correlate of protection against EBOV disease ^20^, it remains unknown whether BDBV-cross-reactive antibodies are sufficient to protect humans against BDBV disease. T-cell responses may provide a complementary layer of heterologous immunity by recognizing multiple, often conserved, linear epitopes and may therefore retain broader activity across related viruses. The cytotoxic BDBV- and SUDV-reactive CD8⁺ T-cell responses identified here, particularly in EVD survivors, may thus represent a relevant component of heterologous immunity. However, neither these cellular responses nor the accompanying binding antibodies can currently be considered correlates of protection against BDBV.

Natural EBOV infection and vaccination produced a similar overall hierarchy of T-cell recognition, with the strongest responses directed against EBOV, substantial recognition of BDBV and SUDV, and limited recognition of MARV. However, survivors exhibited higher response magnitudes, particularly among cytotoxic CD8⁺ T cells, several years after infection. This may reflect differences in antigen load, duration of exposure and the inflammatory environment associated with acute EBOV infection compared with vaccination. Natural infection may also recruit a broader repertoire of T-cell clonotypes recognizing conserved GP epitopes, thereby increasing the magnitude and breadth of long-term memory. Nevertheless, because survivors and vaccinees differed in their modes of antigen exposure and sampling times, these observations should not be interpreted as a direct quantitative comparison between infection and vaccination.

The observed hierarchy of heterologous recognition is broadly consistent with the phylogenetic relatedness of the filovirus glycoproteins. BDBV and SUDV belong to the genus *Orthoebolavirus*, whereas MARV belongs to the distinct genus *Orthomarburgvirus*, which may account for the markedly weaker MARV-directed responses. The limited MARV recognition observed after rVSVΔG-ZEBOV-GP vaccination and in EVD survivors therefore reinforces the specificity of the broader responses detected against BDBV and SUDV. Responses following MVA-BN-Filo require additional caution because this multivalent vector directly encodes the glycoproteins of EBOV, SUDV and MARV. Consequently, SUDV- and MARV-specific responses detected after MVA-BN-Filo administration cannot be attributed exclusively to cross-reactivity. In contrast, BDBV GP is not encoded by this vaccine, making BDBV recognition the clearest measure of heterologous immunity in this group. The limited MARV responses observed despite the inclusion of MARV GP in MVA-BN-Filo may reflect differences in antigen immunogenicity, immunodominance or memory expansion, indicating that sequence-relatedness alone does not fully determine the breadth of the response.

This study has several limitations. The small sample size and exploratory design did not provide sufficient statistical power for formal comparisons between vaccine groups and EVD survivors. Antigen-specific in vitro expansion facilitated the detection of low-frequency memory T-cell responses but did not allow their original ex vivo frequencies to be determined and may have selectively expanded particular T-cell populations. Moreover, responses to complete peptide pools demonstrate heterologous recognition at the T-cell population level but do not establish cross-reactivity of individual T-cell clonotypes. The antibody analyses were limited to binding responses and did not assess neutralizing or protective activity against BDBV. Finally, the detection of cross-reactive immune responses does not demonstrate protection against BDBV.

Nevertheless, our findings demonstrate that EBOV vaccination and natural infection are associated with memory T-cell recognition of BDBV and SUDV, alongside antibody binding to BDBV. Among vaccinees, the positive associations between EBOV- and BDBV-reactive responses within both the cellular and humoral compartments further link the magnitude of EBOV-specific immunity to that of heterologous BDBV recognition. Together with evidence of partial heterologous protection in non-human primates and recent human studies reporting BDBV-reactive antibodies and T cells following rVSVΔG-ZEBOV-GP vaccination, these findings provide an immunological rationale for evaluating existing EBOV vaccines during the ongoing BDBV outbreak.

## Contributors

Conceptualization: AW and YL. Methodology: AW, EL and AA. Investigation: AW, AA, EF, SK, GT. Resources: AW, AA, EL, SF. Data curation: AW, AA. Formal analysis: AW, AA. Visualization: AW, AA. Writing original draft: AW and YL. Writing review and editing: all authors. Supervision: AW and YL. Project administration: AW and YL. Funding acquisition: YL. All authors reviewed and approved the final version of the manuscript.

## Data sharing

The pseudonymised participant data underlying this study cannot be made publicly available because they include sensitive clinical and immunological information and are subject to participant privacy, consent, legal, and ethical restrictions. PREVAC metadata are available in the Recherche Data Gouv repository (DOI: 10.57745/PVXRMC). Requests for access to pseudonymised PREVAC/PREVAC-UP clinical and immunological data should be directed to and will be reviewed by the study sponsors and the principal investigators in the relevant participating countries to ensure compliance with applicable legal, ethical, and regulatory requirements. If access is approved, a data transfer agreement must be established before data transfer. Requests for access to Postebogui data should be directed to the corresponding author and will be considered in accordance with the cohort’s governance procedures and applicable legal and ethical requirements.

## Data Availability

PREVAC metadata are available in the Recherche Data Gouv repository (DOI: 10.57745/PVXRMC). Requests for access to Postebogui data should be directed to the corresponding author

## Acknowledgements

We thank the Ebola virus disease survivors enrolled in the Postebogui cohort and all participants in the PREVAC trial. We also thank the Postebogui and PREVAC study teams for their contribution to these studies.

## References

1. https://sante.gouv.cd/epidemie/ebola-bundibugyo-2026.

2. Kuppalli K, Lokudu R, Azman AS, et al. Reconsidering the rVSVDeltaG-ZEBOV-GP vaccine during the 2026 Bundibugyo virus outbreak. Lancet Infect Dis 2026; 26(9): 875–7.

3. Falzarano D, Feldmann F, Grolla A, et al. Single immunization with a monovalent vesicular stomatitis virus-based vaccine protects nonhuman primates against heterologous challenge with Bundibugyo ebolavirus. J Infect Dis.; 2011: S1082–9.

4. Mire CE, Geisbert JB, Marzi A, Agans KN, Feldmann H, Geisbert TW. Vesicular stomatitis virus-based vaccines protect nonhuman primates against Bundibugyo ebolavirus. PLoS Negl Trop Dis.; 2013: e2600.

5. Woolsey C, Borisevich V, Agans KN, Fenton KA, Cross RW, Geisbert TW. Bundibugyo ebolavirus Survival Is Associated with Early Activation of Adaptive Immunity and Reduced Myeloid-Derived Suppressor Cell Signaling. mBio.; 2021: e0151721.

6. Wiedemann A, Foucat E, Hocini H, et al. Long-lasting severe immune dysfunction in Ebola virus disease survivors. Nat Commun.; 2020: 3730.

7. Wiedemann A, Lhomme E, Huchon M, et al. Long-term cellular immunity of vaccines for Zaire Ebola Virus Diseases. Nat Commun.; 2024: 7666.

8. Felgner J, Clarke E, Hernandez-Davies JE, et al. Broad antibody and T cell responses to Ebola, Sudan, and Bundibugyo ebolaviruses using mono-and multi-valent adjuvanted glycoprotein vaccines. Antiviral Res 2024; 225: 105851.

9. Mdluli T, Wollen-Roberts S, Merbah M, et al. Ebola virus vaccination elicits Ebola virus-specific immune responses without substantial cross-reactivity to other filoviruses. Sci Transl Med 2025; 17(792): eadq2496.

10. Lhomme E, Wiedemann A, Ayouba A, et al. Cross-Reactive Bundibugyo Antibody Responses after Receipt of Licensed Ebola Vaccines. N Engl J Med 2026; 395(6): 612–5.

11. Mayer L, Londt R, Gruener E, Ly ML, Addo MM, Geldmacher C. Cross-reactive T-cell responses against BDBV in humans after rVSVDeltaG-ZEBOV-GP vaccination. Lancet Infect Dis 2026.

12. Badio M, Lhomme E, Kieh M, et al. Partnership for Research on Ebola VACcination (PREVAC): protocol of a randomized, double-blind, placebo-controlled phase 2 clinical trial evaluating three vaccine strategies against Ebola in healthy volunteers in four West African countries. Trials.; 2021: 86.

13. Etard JF, Sow MS, Leroy S, et al. Multidisciplinary assessment of post-Ebola sequelae in Guinea (Postebogui): an observational cohort study. Lancet Infect Dis 2017; 17(5): 545–52.

14. Msellati P, Toure A, Sow MS, et al. [Revival after Ebola: multidisciplinary assessment at 1 year, prospect and follow-up study of surviving patients from Ebola in Guinea (PostEboGui cohort)]. Bull Soc Pathol Exot.; 2016: 236–43.

15. Sow MS, Etard JF, Baize S, et al. New Evidence of Long-lasting Persistence of Ebola Virus Genetic Material in Semen of Survivors. J Infect Dis.; 2016: 1475–6.

16. Team PS, Kieh M, Richert L, et al. Randomized Trial of Vaccines for Zaire Ebola Virus Disease. N Engl J Med 2022; 387(26): 2411–24.

17. Ayouba A, Toure A, Butel C, et al. Development of a Sensitive and Specific Serological Assay Based on Luminex Technology for Detection of Antibodies to Zaire Ebola Virus. J Clin Microbiol.; 2017: 165–76.

18. Sullivan NJ, Hensley L, Asiedu C, et al. CD8+ cellular immunity mediates rAd5 vaccine protection against Ebola virus infection of nonhuman primates. Nat Med.; 2011: 1128–31.

19. Marzi A, Engelmann F, Feldmann F, et al. Antibodies are necessary for rVSV/ZEBOV-GP-mediated protection against lethal Ebola virus challenge in nonhuman primates. Proc Natl Acad Sci U S A.; 2013: 1893–8.

20. Roozendaal R, Hendriks J, van Effelterre T, et al. Nonhuman primate to human immunobridging to infer the protective effect of an Ebola virus vaccine candidate. NPJ Vaccines.; 2020: 112.

