## Supplementary material for "Cross-reactive antibody and functional T-cell responses to Bundibugyo virus induced by Zaire ebolavirus vaccination and infection": Supplemenatary information

Wiedemann A. *et al*

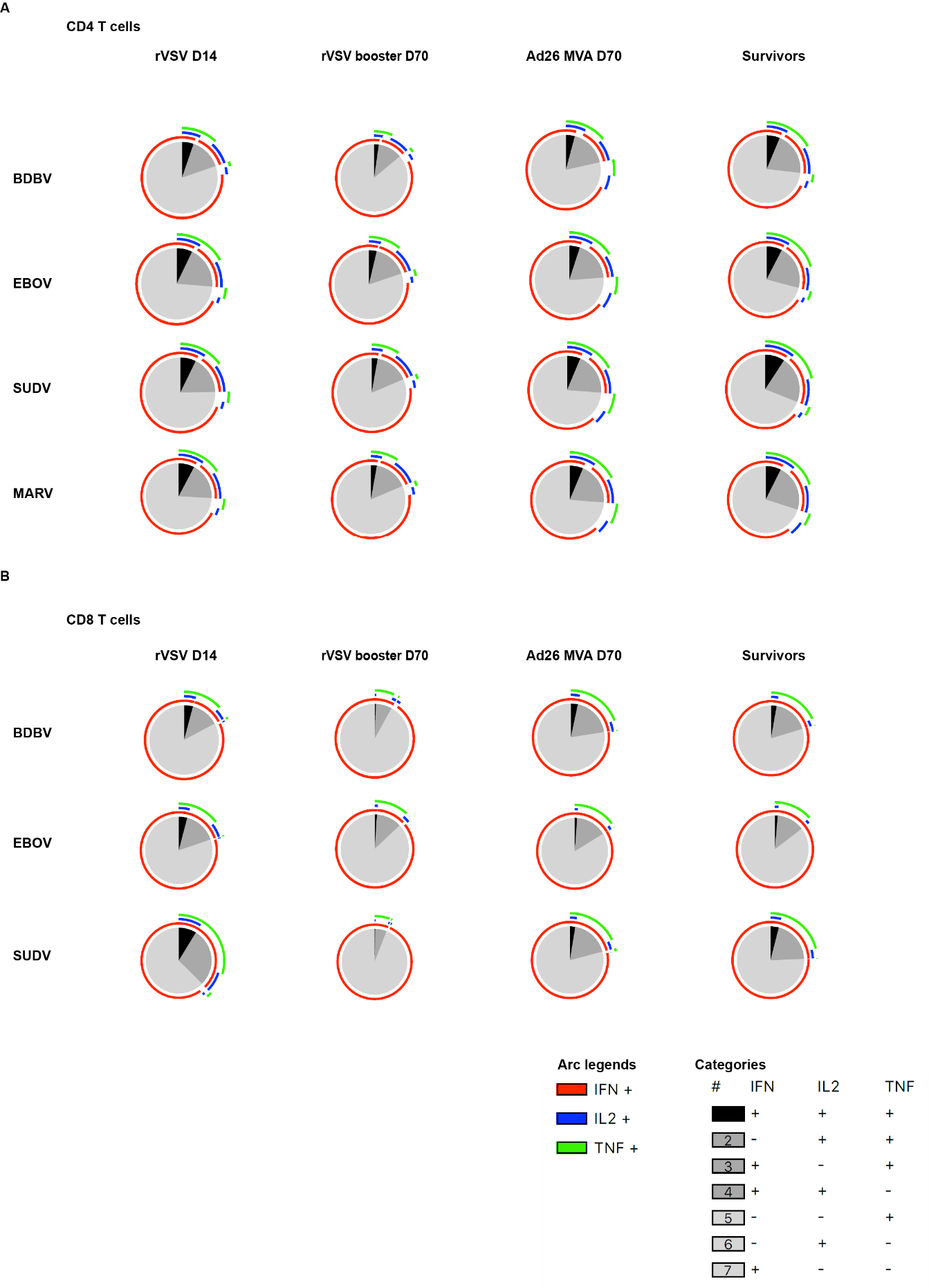

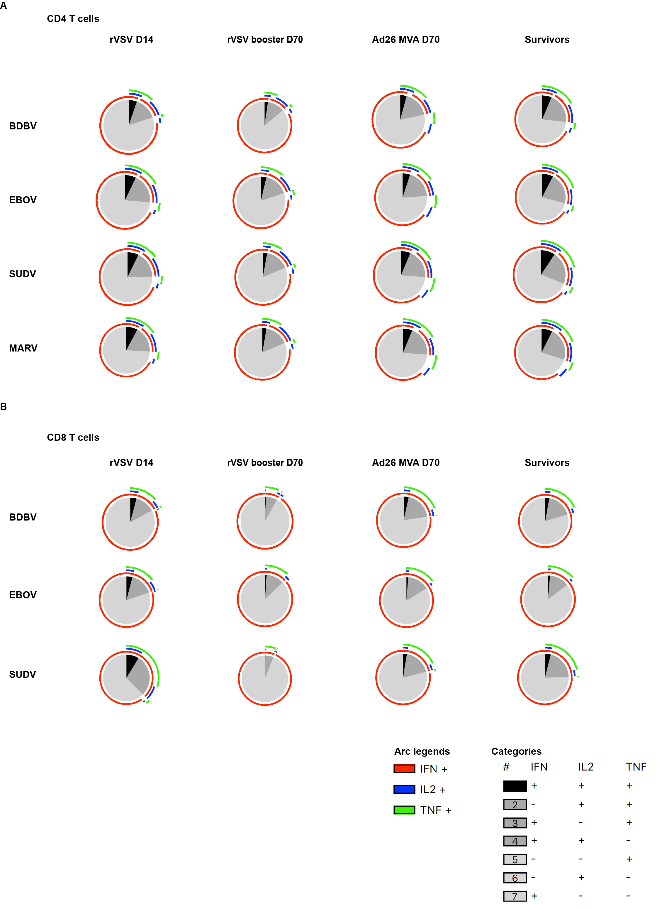

**Supplementary Figure 1. Functional profiles of filovirus GP-specific CD4⁺ and CD8⁺ T-cell responses in vaccinated participants and EVD survivors**. Polyfunctionality of GP-specific CD4⁺ (A) and CD8⁺ (B) T-cell responses was assessed by Boolean analysis of IFN-γ, IL-2 and TNF expression following antigen-specific in vitro expansion and stimulation with overlapping peptide pools spanning the glycoproteins of EBOV, BDBV, SUDV or MARV. Analyses are shown for participants 14 days after a single dose of rVSVΔG-ZEBOV-GP, 14 days after a second dose of rVSVΔG-ZEBOV-GP, 14 days after MVA-BN-Filo boosting of Ad26.ZEBOV-primed participants, and for EVD survivors approximately 3 years after discharge from an Ebola treatment centre. Pie charts depict the distribution of functional subsets according to the combination of cytokines produced. Arcs indicate the frequency of cells expressing IFN-γ, IL-2 and TNF within Filovirus-specific CD8⁺ and CD4⁺ T-cell responses. Only participants meeting the predefined response criterion (an antigen-stimulated response at least threefold higher than the corresponding unstimulated background) were included. MARV-specific CD8⁺ T-cell responses are not shown because too few participants in each group met this criterion to allow reliable SPICE analysis. Data were analysed and displayed using SPICE version 5.22.

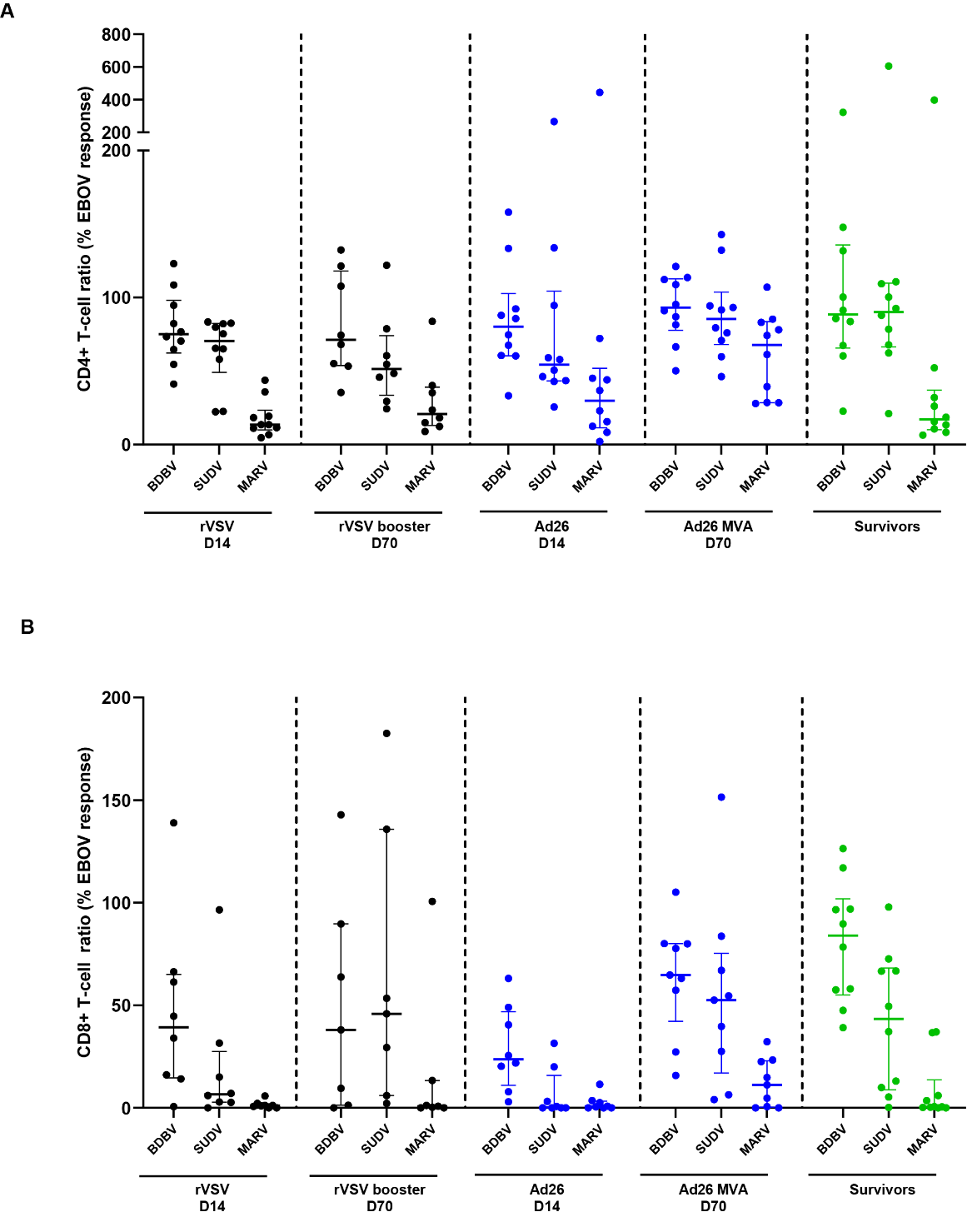

**Supplementary Figure 2. Relative breadth of filovirus-specific CD4⁺ and CD8⁺ T-cell responses.** Relative (A) CD4⁺ and (B) CD8⁺ T-cell responses to BDBV, SUDV and MARV were expressed as percentages of the corresponding EBOV-specific response. For each participant, the unstimulated response was subtracted from each antigen-stimulated response, and the resulting background-subtracted heterologous response was divided by the background-subtracted EBOV response and multiplied by 100. Ratios were calculated only for participants with a positive EBOV-specific response, defined as an antigen-stimulated response at least threefold higher than the corresponding unstimulated response. Negative background-subtracted responses to heterologous peptide pools were set to zero. Ratios were not capped at 100%, as heterologous responses could exceed the corresponding EBOV response. Each symbol represents an individual participant. Horizontal lines and error bars indicate the median and interquartile range. Of note, because MVA-BN-Filo encodes the glycoproteins of EBOV, SUDV and MARV, SUDV- and MARV-to-EBOV ratios at D70 in the Ad26.ZEBOV/MVA-BN-Filo group represent relative response magnitude and cannot be attributed to cross-reactivity alone.

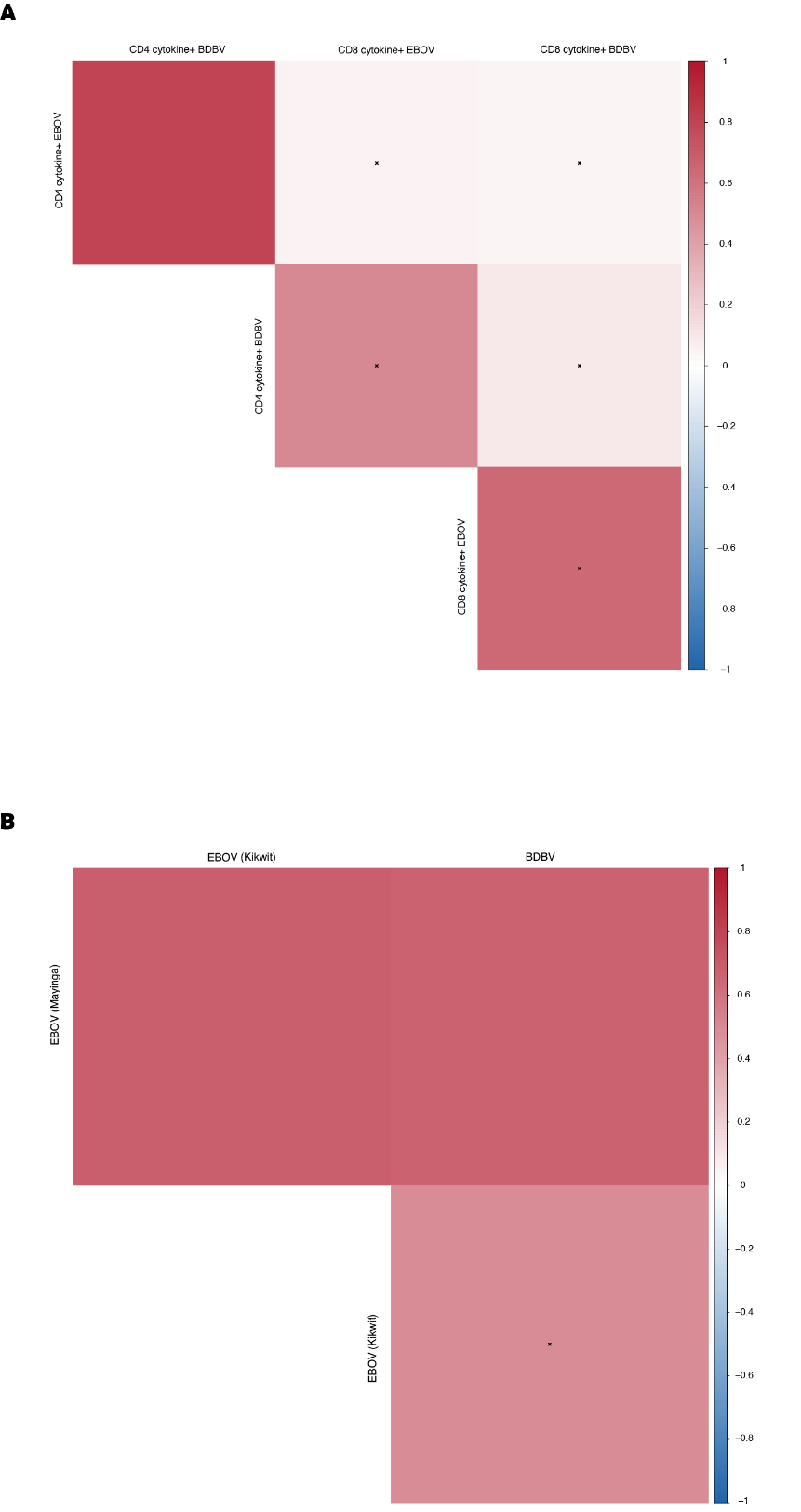

**Supplementary Figure 3. Correlations between EBOV- and BDBV-reactive immune responses in EVD survivors**. (A) Spearman correlation matrix of total cytokine-producing CD4⁺ and CD8⁺ T-cell responses to EBOV and BDBV GP peptide pools. Total cytokine-producing cells were defined as cells expressing IFN-γ, IL-2 and/or TNF. (B) Spearman correlation matrix of antibody-binding responses to EBOV Mayinga, EBOV Kikwit and BDBV glycoproteins. Colours indicate Spearman correlation coefficients, ranging from −1 (blue) to +1 (red). Crosses indicate correlations that were not statistically significant after adjustment for multiple testing (adjusted P ≥ 0.05). Analyses were performed on n=10 EVD survivors.

**Postebogui Study Group**

Ahidjo Ayouba (IRDUMI233-INSERMU1175-Montpellier University, Montpellier, France), Sylvain Baize (Pasteur Institute/Unit of Biology of Emerging Viral Infections, Lyon, France), Kaba Bangoura (Donka National Hospital, Conakry, Guinea), Alimou Barry (IRDUMI233-INSERMU1175-Montpellier University, Montpellier, France), Moumié Barry (Donka National Hospital, Conakry, Guinea), Mamoudou Cissé (Forecariah Prefectoral Hospital, Forecariah, Guinea), Mohammed Cissé (Donka National Hospital, Conakry, Guinea), Eric Delaporte (IRDUMI233-INSERMU1175-Montpellier University, Montpellier, France), Jean-François Delfraissy (Reacting, INSERM, Paris, France), Christelle Delmas (Reacting, INSERM, Paris, France), Alice Desclaux (IRDUMI233-INSERMU1175-Montpellier University, Montpellier, France), Saliou Bella Diallo (Donka National Hospital, Conakry, Guinea), Mamadou Safiatou Diallo (Donka National Hospital, Conakry, Guinea), Mariama Sadjo Diallo (Donka National Hospital, Conakry, Guinea), Jean François Étard (IRDUMI233- INSERMU1175-Montpellier University, Montpellier, France), Cécile Etienne (Reacting, INSERM, Paris, France), Ousmane Faye (Donka National Hospital, Conakry, Guinea), Ibrahima Fofana (Macenta Prefectoral Hospital, Macenta, Guinea), Bruno Granouillac (IRDUMI233-INSERMU1175-Montpellier University, Montpellier, France), Esther Hereth Hébert (IRDUMI233-INSERMU1175- Montpellier University, Montpellier, France), Suzanne Izard (IRDUMI233-INSERMU1175-Montpellier University, Montpellier, France), Djenaba Kassé (Donka National Hospital, Conakry, Guinea), Alpha Kabinet Keita (IRDUMI233-INSERMU1175-Montpellier University, Montpellier, France), Sakoba Keita (Ministry of Health, Conakry, Guinea), Lamine Koivogui (Institut National de la Santé Publique, Conakry, Guinea), Cécé Kpamou (IRDUMI233- INSERMU1175-Montpellier University, Montpellier, France), Christine Lacabaratz (INSERM, Paris, France), Sandrine Leroy (IRDUMI233-INSERMU1175-Montpellier University, Montpellier, France), Claire Levy Marchal (INSERM, Paris, France), Yves Levy (INSERM, Paris, France), N’Fally Magassouba (Laboratory of Virology, Conakry University, Projet de Recherche sur les Fièvres Hémorragiques en Guinée, Conakry, Guinea), Laura March (IRDUMI233-INSERMU1175-Montpellier University, Montpellier, France), Vincent Mendiboure (ALIMA, Conakry,Guinea), Philippe Msellati (IRDUMI233-INSERMU1175-Montpellier University, Montpellier, France), Harissatou Niane (Donka National Hospital, Conakry, Guinea), Martine Peeters (IRDUMI233-INSERMU1175-Montpellier University, Montpellier, France), Yves-Marie Pers (IRDUMI233-INSERMU1175-Montpellier University, Montpellier, France), Hervé Raoul (Laboratoire P4 Inserm-Jean Mérieux, US003 Inserm, Lyon, France), Sidi Lamine Sacko (Macenta Prefectoral Hospital, Macenta, Guinea), Ibrahima Savané (Macenta Prefectoral Hospital, Macenta, Guinea), Mamadou Saliou Sow (Donka National Hospital, Conakry, Guinea), Bernard Taverne (IRDUMI233-INSERMU1175-Montpellier University, Montpellier, France), Abdoulaye Touré (Department of Public Health, Conakry University, Conakry, Guinea), Fodé Amara Traoré (Donka National Hospital, Conakry, Guinea), Falaye Traoré (Institut National de la Santé Publique, Conakry, Guinea), Yamoussa Youla (N’Zérékoré Regional Hospital, N’Zérékoré, Guinea), Yazdan Yazdanpanah (Reacting, INSERM, Paris, France)

**PREVAC Study Team**

Jamila Aboulhab2 M.D., Michelle Aguirre-MacKenzie12 B.S., Pauline Akoo3 M.B., Ch.B., Esther Akpa2 M.S.N., M.P.H., R.N., Robert Akpata1 M.D., Sara Albert17 B.A., M.P.H, Boni Maxime Ale4 M.D., M.Sc., M.P.H., Serry Alimamy-Bangura11 M.B., Ch.B., Benetta C. Andrews10 M.D., Stephane Anoma6 M.D., Negin Atri2 M.P.H., C.P.H., Augustin Augier6 M.Com., Ken Awuondo3 M.Sc., Ahidjo Ayouba30 Ph.D., Moses Badio10 M.Sc., Aminata Bagayoko1 M.D., Abby Balde17 M.P.H., Joséphine Balssa1,8 Pharm.D., Lamin Molecule Bangura11 B.Sc., Kesha Barrington17 M.P.A., Eric Barte de Saint Fare6 B.A., Beth Baseler17 M.S., Ali Bauder12 B.A., P.M.P., Claire Bauduin4 M.Sc, Luke Bawo10 M.Sc., Abdoul Habib Beavogui9 M.D., Ph.D., Michael Belson2 B.S., Safaa Ben-Farhat4 M.Eng., Marion Bererd6 B.S., Nicolas Bernaud1,18 M.Sc., Teedoh Beyslow10 Pharm.D., Neirade Biai30 M.Sc., Jeanne Billioux2 M.D., Shere Billouin-Frazier17 M.Sc., Blandine Binachon4 M.D., M.P.H., Julie Blie10 M.Sc., Patricia Boison17 M.S., Fatorma Bolay10 Ph.D., Aliou Boly6 M.M., Rachael Elizabeth Bonawitz12 M.D., M.S., Anne-Gaëlle Borg6 M.C.M., Samuel Bosompem2 Pharm.D. M.Sc., Courtney Bozman28 M.Sc., Tyler Brady2 M.P.H., Sarah Browne10 R.N., B.S.N., Ryan Bullis12 Ph.D., Gabrielle Caberia1 Pharm.D. candidate, Barbara Cagniard1 Ph.D., Kelly Cahill2 R.N, M.Sc., C.C.R.C., R.A.C., Yingyun Cai28 Ph.D., Aissata Abdoulaye Camara6 M.Sc., Aboubacar Keira Camara1 M.D., M.Sc., Alseny Modet Camara6 M.D., Cécilia Campion4 M.Sc., Alexandre Cantan4,29 B.Sc., Jennifer Cash17 B.S., Michael Chea10 B.Sc., Geneviève Chêne4 M.D., Ph.D., Edward Choi3 Ph.D., Michelle Chouinard6 M.S.W., Florence Chung1 Ph.D., Lucy Chung2 Pharm.D., Papa Ndiaga Cisse14 Ph.D., Elfrida Cline-Cole17 M.A., Céline Colin4 M.Sc., Beth-Ann Coller12 Ph.D., Djélikan Siaka Conde1 M.D., Katherine Cone2 M.S.W., LCSW-C, C-SWHC, Laurie Connor12 M.S., Nicholas Connor3 M.Sc., Joseph Boye Cooper10 M.Sc., Sandrine Couffin-Cadiergues1 Ph.D., Mariam Coulibaly20 Pharm.D., M.Sc., Page Crew2 Pharm.D., M.P.H, B.C.P.S., Sandrine Dabakuyo-Yonli4 Pharm.D., Ph.D., Djeneba Dabitao20 Pharm.D., Ph.D., Bionca Davis5 M.P.H., Gibrilla Fadlu Deen11 M.D., M.Sc., Jean-François Delfraissy1 M.D., Ph.D., Christelle Delmas1 M.Sc., Mahamadou Diakite20 Pharm.D., D.Phil, Alpha Diallo1,8 M.D., M.P.H., Fatoumata Abdoulaye Diallo6, M.D., Mamadou Saliou Diallo6 M.D., M.P.H., Ayouba Diarra20 M.Sc., Samba Diarra20 M.Sc., Ph.D., Oualy Diawara19 B.S., Ilo Dicko 20 M.D, M.P.H., Bonnie Dighero-Kemp28 B.Sc., Samba Diop21 M.Sc., Ph.D., Waly Diouf14 Ph.D., Saurabh Dixit2 Ph.D., Barry Djenabou6 M.Sc., Laurie Doepel2 B.A., Eric D'Ortenzio1,7,8 M.D., M.P.H., Seydou Doumbia20 M.D., Ph.D., Moussa Moise Doumbia19 M.D., Nelson Dozier28 M.Sc., Natasha Dubois Cauwelaert1,8 Ph.D., Alain DuChêne5 B.S., Michael Duvenhage17 N.DIP.IT., Risa Eckes2 R.N., Elizabeth Elliott2 M.Sc., Luisa Enria3 Ph.D., Hélène Espérou1 M.D., Cécile Etienne1 M.Sc., Allison Eyler17 H.S.D., Lawrence Fakoli10 M.Sc., Mosoka Fallah10 Ph.D., Marie-Alix Fauvel1 M.Sc., Sylvain Faye14 Ph.D., John Fayiah10 M.Sc., Suzanne Fleck3 Ph.D., Vemy Fofana6 B.Comp, Karine Fouth Tchos2 M.D., M.P.H., Kokulo Franklin10 M.Phil., M.Sc., Daniela Fusco1 Ph.D., Auguste Gaddah13 Ph.D., Marylène Gaignet1 M.Sc., Katherine Gallagher3 Ph.D., Julie Gardner12, B.S., Harrison Gichini28 M.Sc., Julia Garcia Gozalbes1 M.D., Greg Grandits5 M.S, Maima Gray10 B.Pharm., Brian Greenwood3 M.D., Robin Gross28 M.Sc., Louis Grue17 R.N., B.S., B.S.N., Birgit Grund27 Ph.D., Oumar Guindo20 M.Sc., Pharm.D., Swati Gupta12 Dr.P.H., M.P.H., Fadima Haidara19 M.D., Benjamin Hamzé1 Pharm.D., Emma Hancox3 M.Sc., Patricia Hensley3 M.Ph., Lisa Hensley28 Ph.D., M.S.P.H., Betsey Herpin2, M.S.N, Elisabeth Higgs2 M.D., D.T.M.H., M.I.A., Trudi Hilton3 B.Pharm., M.Sc., Mickael Hneino1, Ph.D., Tracey-Ann Hoeltermann28, B.Sc., M.P.H., , Horace Preston Holley17 M.D., Marie Hoover16 Ph.D., Natasha Howard3 Ph.D., Melissa Hughes12 B.A., M.B.A., C.P.M., P.M.P., Sesay Idrissa32 B.Sc, Skip Irvine12 B.S., David Ishola3 M.D., Ph.D., Yvonne Jato2 M.P.H., Madison Joe10 M.Sc., Melvin Johnson10 M.Sc., Aboubacar Sidiki Kaba6 M.D., Jonathan Kagan2 Ph.D., Kade Kallon17, M.Sc., Michael Kamara3 MB.ChB, M.Sc., Myriam Kante4 B.S., Judith Katoudi6 M.D., M.P.H., Cheick Mohamed Keita6 M.D., Sakoba Keita15 M.D., Seykou Keita22 M.D., Stephen B. Kennedy10M.D., M.Sc., Babajide Keshinro13 M.B.B.S., F.W.A.C.P., Hassan Kiawu10 M.Sc., Mark Kieh10 M.D., M.S.M.H.C., Brent Killinger12 B.A., Moumouni Kinda6 M.D., M.B.A., Matthew Kirchoff2 Pharm.D., M.Sc., M.B.A., Gregory Kocher28 M.Sc., Mamoudou Kodio19 Pharm.D., Brian Kohn3 B.Sc., Lamine Koivogui23 Pharm.D., Ph.D., Richard Kojan6 M.D., Cece Francis Kolié6 Pharm.D., Jacques Seraphin Kolié6 M.D., David Kollie10 B.Sc., Stacy Kopka17 M.S., Bockarie Koroma11 B.Pharm., Dickens Kowuor3 B.Sc., M.Sc., Ph.D., Catherine Kpayieli-Freeman10 M.Sc., Ange-Marie D. Kpetigo31 M.Sc., Christine Lacabaratz1,18 Ph.D., Boris Lacarra1 M.D., Laurie Lambert17 B.S., Courtney Lambeth12 B.S., Solange Lancrey-javal1,8 Pharm.D., H. Clifford Lane2 M.D., Shadrach Langba10 B.Sc., Bolarinde Lawal3 M.Sc., Andrew Wen-Tseng Lee12 M.D., Shona Lee3 Ph.D., Shelley Lees3 Ph.D., Annabelle Lefevre1 M.D., Bailah Leigh11 M.D., M.Sc., Frederic Lemarcis1 Ph.D., Yves Lévy1,18 M.D., Ph.D., Edouard Lhomme26 M.D., Ph.D., Janie Liang28 M.Sc., Mameni Linga10 M.Sc., Ken Liu12 Ph.D., Brett Lowe3 M.Phil., Julia Lysander10 M.Sc., Ibrah Mahamadou6 Pharm.D., Irina Maljkovic-Berry28 Ph.D., Marvington Mambiah10 A.Sc., Daniela Manno3 M.D., Ph.D., Jonathan Marchand2,17 M.S., Lindsay Marron28 M.Sc., Moses B.F. Massaquoi10 M.D., M.Sc., , Charly Matard4 B.S., Steven Mazur28 B.S., John McCullough16 B.S., Chelsea McLean13 Ph.D., Noémie Mercier1 Pharm.D., Pauline Michavila6 B.Bus., Tracey Miller17 R.N., B.S.N., Niouma Pascal Millimouno 6 M.D., Alejandra Miranda17 M.S., Soumaya Mohamed6 B.J., Tom Mooney3 B.A., Dally Muamba6 M.D., James Mulbah11 B.Pharm., Rita Lukoo Ndamenyaa6 M.D., M.Sc., James Neaton5 Ph.D., Désiré Neboua1 M.D., Micki Nelson12 B.S.N., M.S., Kevin Newell17 M.P.H., M.Ed., Vinh-kim Nguyen24 M.D., Yusupha Njie3 B.Sc., Wissedi Njoh17 M.S.N., Matthew Onorato12 B.S., Uma Onwuchekwa22 B.Sc., Susan Orsega2 M.S.N., FNP-BC, Inmaculada Ortega-Perez1,8 Ph.D., M.P.H., Cynthia Osborne17 B.S., Tuda Otieno3 M.SC., Davy Oulaï4 M.D., Sushma Patel12 M.S., P.M.P., Danielle Peart2 B.S., Martine Peeters30 Ph.D., James Pettitt28 M.Sc., Nathan Peiffer-Smadja1 M.D., Ph.D., Robert Phillips3 M.Sc., Jerome Pierson2 Ph.D., Peter Piot3 M.D., Ph.D., Micheal Piziali2 J.D., M.Sc., Stéphany Pong1,8 Pharm.D., Elena Postnikova2 Ph.D., Dudley Pratt3 M.D., Calvin Proffitt17 M.A., Alexandre Quach1 M.D., Sinead Quigley1 M.S.Sc., Nadeeka Randunu2,17 B.Sc., M.B.A., Laura Richert26 M.D., Ph.D., Priscille Rivière1 M.Sc., Céline Roy4,29, Ph.D., Amy Falk Russell12 M.S., Philip Sahr10 M.D., Katy Saliba2, M.Sc., Ph.D., Mohamed Samai11 M.B.B.S., Ph.D., Sibiry Samake20 Pharm.D., M.Sc., Jen Sandrus17 A.A., Ibrahim Sanogo20 Ms.P., M.D., Yeya Sadio Sarro20 Pharm.D., Ph.D., Serge Sawadogo6 M.D., M.Sc., Sani Sayadi6 M.D., M.P.H., Maxime Schvartz1 M.D., Christine Schwimmer4 Ph.D., Fatou Secka3 B.Sc., M.Sc., MB.ChB, Heema Sharma28M.Sc., Denise Shelley17 M.S., Bode Shobayo10 M.Sc., Sophia Siddiqui2 M.D., M.P.H., Jakub Simon12 M.D., Shelly Simpson17 M.S., Billy Muyisa Sivahera6 M.D., , Mary Smolskis2 B.S.N., M.A., Elizabeth Smout3 M.D., M.Sc., Emily Snowden3 M.A., Anne-Aygline Soutthiphong4,29 M.Sc., Amadou Sow6 M.Sc., Samba O. Sow22 M.D., M.Sc., Ydrissa Sow2 M.D., M.P.H., Michael Stirratt25 Ph.D., Léa Surugue1 M.J., Sienneh Tamba10 R.N., B.S.N., Cheick Tangara20 M.Sc., Milagritos D. Tapia22 M.D., Julius Teahton10 M.Sc., Jemee Tegli10 M.Sc., Monique Termote4 M.Sc., Guillaume Thaurignac30 M.Sc., Rodolphe Thiebaut4 M.D., Ph.D., Greg Thompson5 B.S., John Tierney2 B.S.N., M.P.M., Daniel Tindanbil3 M.Sc., Abdoulaye Touré23 Pharm.D., M.P.H., Ph.D., Elvis Towalid10 B.Pharm, Stacey Traina12 B.S., Awa Traore19 Pharm.D., Tijili Tyee10 Pharm.D., David Vallée1 Pharm.D., Renaud Vatrinet1 Ph.D., Corine Vincent4 M.Sc., Susan Vogel2 R.N, B.S.N., Cedrick Wallet4 M.Sc., Travis Warren2 Ph.D., Deborah Watson-Jones3 M.D., Ph.D., Wade Weaver28 M.Sc., Deborah Wentworth5 M.P.H., Cecelia Wesseh10 B.Sc., Hilary Whitworth3 Ph.D., Jimmy Whitworth3 Ph.D., Aurelie Wiedemann1,18 Ph.D., Wouter Willems13 Ph.D, Barthalomew Wilson10 M.Sc., Jayanthi Wolf12 Ph.D., Alie Wurie11 M.D., M.Sc., Delphine Yamadjako17 M.S., Marcel Yaradouno6 M.Sc., Quiawiah Yarmie10 M.Sc., Yazdan Yazdanpanah1,7,8 M.D., Ph.D., Shuiqing Yu28 B.S., Zara Zeggani6 M.Sc., Huanying Zhou28 B.S.

Affiliations

1 French Institute for Health and Medical Research (Inserm), 75013 Paris, France

2 National Institute of Allergy and Infectious Diseases, Bethesda, MD, USA or under contract/subcontract to NIAID

3 London School of Hygiene & Tropical Medicine, London, UK

4 Univ. Bordeaux, INSERM, Institut Bergonié, CHU de Bordeaux, CIC-EC 1401, Euclid/F-CRIN Clinical trials platform, F-33000 Bordeaux, France

5 School of Public Health, University of Minnesota, Minneapolis, MN, USA

6 The Alliance for International Medical Action, Alima, B.P.15530 Dakar, Sénégal

7 AP-HP, Hôpital Bichat-Claude Bernard, Service de Maladies Infectieuses et Tropicales, Paris F-75018, France

8 ANRS Emerging Infectious Diseases, Paris, France

9 Centre National de Formation et de Recherche en Santé Rurale de Maferinyah, Maferinyah, Guinea

10 Partnership for Research on Ebola Virus in Liberia (PREVAIL), Monrovia, Liberia

11 College of Medicine and Allied Health Sciences (COMAHS), University of Sierra Leone, Freetown, Sierra Leone

12 Merck Sharp & Dohme Corp, Inc., Kenilworth, NJ, USA

13 Janssen Vaccines and Prevention BV Leiden, The Netherlands

14 Département de Sociologie, FLSH, Université Cheikh Anta DIOP, Dakar, Sénégal

15 Agence Nationale de Sécurité Sanitaire, Conakry, Guinea

16 Advanced BioMedical Laboratories, L.L.C., 1605 Industrial Hwy, Cinnaminson, NJ, USA

17 Clinical Monitoring Research Program Directorate, Frederick National Laboratory for Cancer Research.

18 Vaccine Research Institute, Univ. Paris Est Créteil, Henri Mondor Hospital, Créteil, France

19 Centre pour le Développement des Vaccins, Ministère de la Santé, Bamako, Mali

20 University Clinical Research Center (UCRC), University of Sciences, Techniques and Technologies of Bamako (USTTB), Bamako, Mali

21 Liberia Institute for Biomedical Research Ethics Committee/National, Monrovia, Liberia

22 Center for Vaccine Development and Global Health, University of Maryland School of Medicine, 685 West Baltimore Street Baltimore, MD 21201-1509, USA

23 INSP (Institut Nationale de Santé Publique), Conakry, Guinea

24 École de santé publique de l’Université de Montréal, Montréal, Canada

25 National Institute of Mental Health, Bethesda, MD, USA

26 Univ. Bordeaux, INSERM, Institut Bergonié, CHU de Bordeaux, CIC-EC 1401, Euclid/F-CRIN clinical trials platform and U1219 BPH Inria Sistm, F-33000 Bordeaux, France

27 School of Statistics, University of Minnesota, Minneapolis, MN, USA

28 Integrated Research Facility at Fort Detrick (IRF-Frederick), National Institute of Allergy and Infectious Diseases, National Institutes of Health (NIH), Fort Detrick, Frederick, MD, USA

29 Univ. Bordeaux, INSERM, MART, UMS 54, F-33000 Bordeaux, France

30 Recherche Translationnelle Appliquée au VIH et aux Maladies Infectieuses, University of Montpellier, Institut de Recherche pour le Développement, INSERM, 34090, Montpellier, France

31	Bordeaux Population Health Research Centre, Université de Bordeaux, Inserm, and Inria, Bordeaux, France

32	University of Management and Technology UNIMTECH, FreeTown, Sierra Leone.
